# A Numerical Study of the Current COVID-19 Spread Patterns in India, the USA and the World

**DOI:** 10.1101/2020.10.05.20206839

**Authors:** Hemanta K. Baruah

## Abstract

In this article, we are going to study the current COVID-19 spread patterns in India and the United States. We are interested to show how the daily increase in the total number of cases in these two countries is affecting the COVID-19 spread pattern in the World. For the study, we have considered the cumulative total numbers of cases in India, the United States and the World. We have found that the situation in the United States is already on the threshold of a change towards retardation. In the World as a whole also we have observed that a similar conclusion can be made. In India, the situation can be expected to move towards betterment soon, and once that happens the situation in the World as a whole would start improving. We shall demonstrate that as long as the rate of change of the logarithm of the cumulative total number of cases with respect to time in a pandemic continues to reduce, the pattern of growth would continue to remain nearly exponential, and as soon as it is seen that the rate of change starts to become nearly constant the growth can be expected to start to change towards a nearly logarithmic pattern.

## Introduction

A look at the daily increase in the cumulative totals of the COVID-19 cases in India, the United States of America and the World gives us an apparently clear picture how much India is contributing to the daily increase in the total number of cases in the World, and in comparison how much the USA is currently contributing towards that. For example, on October 1 the increase in the World total were nearly 320,000 out of which India alone contributed nearly 81,000 which are about 25% of the increase in the World total, while on that day the USA contributed around 47,000 which are about 15% of the increase in the World total.

In what follows, we are going for a numerical study of the rate at which India is contributing towards the daily increase in the cumulative total number of COVID-19 cases in the world in comparison to the rate at which the USA is contributing. For this we need not go for any compartmental epidemiological model [1. 2, 3]. Indeed, for the World as a whole and for India too, the compartmental models may not probably be very suitable because of geographical and economic heterogeneity of the susceptible population. Further, we are going to study the current situation only for which we do not really need to take resort to the compartmental epidemiological models. In our eyes, unless the data regarding recovered cases and deaths are absolutely reliable, the epidemiological models such as SIR (Susceptible-Infectious-Recovered), SIS (Susceptible-Infectious-Susceptible) and SIRD (Susceptible-Infectious-Recovered-Dead) cannot be criticized. The population exposed to the disease may not have economic homogeneity, and in that situation the SEIR (Susceptible-Exposed-Infectious-Recovered) model similarly cannot be criticized for unacceptable results. Such mathematical models were established under certain assumptions. If those assumptions are not valid it is obvious that the results returned would be unacceptable.

In our present study we shall use data of the cumulative total number of cases only. As a matter of fact, in the initial stage of the pandemic this number was of the population that showed some symptoms of the disease. Later on, when planned testing for COVID-19 positivity got started, those who were tested included people without any symptoms too. So it is obvious that whereas in the initial stage those with symptoms were included in the total number of cases, later on this figure became dependent on the number of tests performed. If due to some reason lesser number of tests was performed in a particular region on any particular day, the increase in the total number of effected cases would automatically be less than that seen on any other day in which more tests were performed. Therefore it has to be agreed upon that the cumulative total number of cases is indeed situation dependent in the sense that the randomness that was inherent in the initial stage of the pandemic is absent in the current number of cases. This may have an adverse effect even in our kind of a study in which only the cumulative total number of cases are being used. Indeed, this would automatically affect the results returned by application of the compartmental epidemiological models too.

We shall use a simple numerical procedure of our own for this study using data from Worldometers.info [4]. Regarding our method of forecasting without using the standard epidemiological methods [5, 6, 7, 8, 9], it was observed that our numerical method does work very well in forecasting of the spread pattern during the nearly exponential phase. In this article, we shall put forward a way to compare the rates of growth of the natural logarithm of the cumulative total of the cases of the pandemic in India, the USA and the World as a whole. Our objective is to demonstrate that as long as the rate of change of the logarithm of the cumulative total number of cases with respect to time in a pandemic continues to reduce, the pattern of growth would continue to remain nearly exponential, and as soon as it is seen that the rate of change starts to become nearly constant the growth would start to change towards a nearly logarithmic pattern.

## Methodology

It is apparent from the graphs published by Worldometers.info [4] that the spread patterns are still approximately exponential in India, in the USA and in the World. Therefore the function

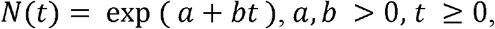

where *N*(*t*) is the cumulative total number of cases at time *t*, should fit the concerned data. Write *z(t) = log*_*e*_ *N*(*t*). We shall now proceed to study the values of Δ*z*(*t*), the first order differences of *z*(*t*).

It was observed [5, 6, 7, 8, 9] that from Δ*z*(*t*) we can extract an important information. When the pandemic continues to grow exponentially, Δ*z*(*t*) would continue to decrease linearly in time. If it continues to be very nearly constant for a sufficiently long duration after continuing to decrease linearly earlier, that should be taken as a signal that the pattern might start to change to a nearly logarithmic one soon after [10]. We shall now show how Δ*z*(*t*) can lead us to conclude about the current situations in India, in the USA and in the World.

## Analysis and Discussions

In Tables-1, 2 and 3, we shall now show the values of *N(t), z(t)* and Δ*z*(*t*) of the World, of the United States of America and of India respectively for 15 days from September 17 to October 1. The data have been taken from the Woldometers.info [4] dated October 2. It should be mentioned here that the data source Woldometers.info gets edited sometimes creating some small changes in the figures thereby. This is why we have cited the source with the date concerned. However, such changes are very insignificant with respect to the largeness of the numbers.

**Table-1:** World: Values of *N(t), z(t)* and Δ*z*(*t*) from 17 September to 1 October.

| Dates | $N(t)$ | $z(t)$ | $\Delta z(t)$ |
| --- | --- | --- | --- |
| Oct. 1 | 34469967 | 17.3556 | 0.009332 |
| Sept. 30 | 34149803 | 17.34627 | 0.009287 |
| Sept. 29 | 33834125 | 17.33698 | 0.008554 |
| Sept. 28 | 33545940 | 17.32843 | 0.00691 |
| Sept. 27 | 33314931 | 17.32152 | 0.00778 |
| Sept. 26 | 33056748 | 17.31374 | 0.009125 |
| Sept. 25 | 32756467 | 17.30461 | 0.009774 |
| Sept. 24 | 32437864 | 17.29484 | 0.009552 |
| Sept. 23 | 32129499 | 17.28529 | 0.009673 |
| Sept. 22 | 31820223 | 17.27561 | 0.008554 |
| Sept. 21 | 31549195 | 17.26706 | 0.0074 |
| Sept. 20 | 31316586 | 17.25966 | 0.008172 |
| Sept. 19 | 31061697 | 17.25149 | 0.009653 |
| Sept. 18 | 30763308 | 17.24183 | 0.010545 |
| Sept. 17 | 30440619 | 17.23129 | 0.010366 |
| Sept. 16 | 30126687 | 17.22092 | --- |

**Table-2:** USA: Values of *N(t), z(t)* and Δ*z*(*t*) from 17 September to 1 October.

| Dates | $N(t)$ | $z(t)$ | $\Delta z(t)$ |
| --- | --- | --- | --- |
| Oct. 1 | 7494671 | 15.8297 | 0.006343 |
| Sept. 30 | 7447282 | 15.82336 | 0.005511 |
| Sept. 29 | 7406353 | 15.81785 | 0.005989 |
| Sept. 28 | 7362126 | 15.81186 | 0.005096 |
| Sept. 27 | 7324706 | 15.80676 | 0.004631 |
| Sept. 26 | 7290867 | 15.80213 | 0.005945 |
| Sept. 25 | 7247651 | 15.79619 | 0.00743 |
| Sept. 24 | 7194003 | 15.78876 | 0.006348 |
| Sept. 23 | 7148483 | 15.78241 | 0.005862 |
| Sept. 22 | 7106700 | 15.77655 | 0.00506 |
| Sept. 21 | 7070834 | 15.77149 | 0.005187 |
| Sept. 20 | 7034253 | 15.7663 | 0.004778 |
| Sept. 19 | 7000725 | 15.76152 | 0.006274 |
| Sept. 18 | 6956937 | 15.75525 | 0.007445 |
| Sept. 17 | 6905332 | 15.7478 | 0.006766 |
| Sept. 16 | 6858767 | 15.74104 | ---- |

**Table-3:** India: Values of *N(t), z(t)* and Δ*z*(*t*) from 17 September to 1 October.

| Dates | $N(t)$ | $z(t)$ | $\Delta z(t)$ |
| --- | --- | --- | --- |
| Oct. 1 | 6391960 | 15.67055 | 0.012863 |
| Sept. 30 | 6310267 | 15.65769 | 0.013842 |
| Sept. 29 | 6223519 | 15.64385 | 0.013019 |
| Sept. 28 | 6143019 | 15.63083 | 0.011406 |
| Sept. 27 | 6073348 | 15.61942 | 0.013722 |
| Sept. 26 | 5990581 | 15.6057 | 0.01497 |
| Sept. 25 | 5901571 | 15.59073 | 0.014588 |
| Sept. 24 | 5816103 | 15.57614 | 0.014883 |
| Sept. 23 | 5730184 | 15.56126 | 0.015776 |
| Sept. 22 | 5640496 | 15.54548 | 0.014355 |
| Sept. 21 | 5560105 | 15.53113 | 0.013488 |
| Sept. 20 | 5485612 | 15.51764 | 0.016058 |
| Sept. 19 | 5398230 | 15.50158 | 0.017332 |
| Sept. 18 | 5305475 | 15.48425 | 0.017644 |
| Sept. 17 | 5212686 | 15.46661 | 0.018743 |
| Sept. 16 | 5115893 | 15.44786 | ---- |

In [5] it was shown that in the World outside China, the values of Δ*z*(*t*) were around 0.13705 from March 21 to March 28, around 0.09287 from March 29 to April 4, around 0.05916 from April 5 to April 11, around 0.0407 from April 12 to April 18, and around 0.03323 from April 19 to April 25. It was clear that within the period from March 21 to April 25, the values of Δ*z*(*t*) were steadily reducing. We have found that the average of Δ*z*(*t*) reduced to 0.008978 during the period from September 17 to October 1.

In [6] it was shown that in the United States, the average of Δ*z*(*t*) was 0.02165 during the period from May 3 to May 8. During the period from September 17 to October 1, it was found to be 0.005911.

In [7] it was shown that in India, the average of Δ*z*(*t*) during May 11 to May 24 was 0.051716, and it came down to 0.045584 during May 25 to May 31. It was seen that [8] during June 1 to June 9 it became 0.04063. When the matter was studied with data for 60 consecutive days [9], from June 23 to July 12, the average value of Δ*z*(*t*) was 0.034576, from July 13 to August 1 it was 0.034458, and from August 2 to August 21 it was 0.026449. So we have seen that as time progressed, the values of Δ*z*(*t*) were reducing. During the period from September 17 to October 1, it was found to be 0.014846.

The values of Δ*z*(*t*) have been depicted in Fig. 1. In the figure, Series-1 represents India, Series-2 represents the USA and Series-3 represents the World. We have not gone for a statistical fit of regression equations of Δ*z*(*t*) on *t* because it looks obvious from the Figure that Δ*z*(*t*) is actually showing a decreasing trend for India, while for the USA and the World the values are very nearly constant. It may further be observed that there may be cycles of length 8 days from one trough to another in all three cases.

**Fig. 1:**
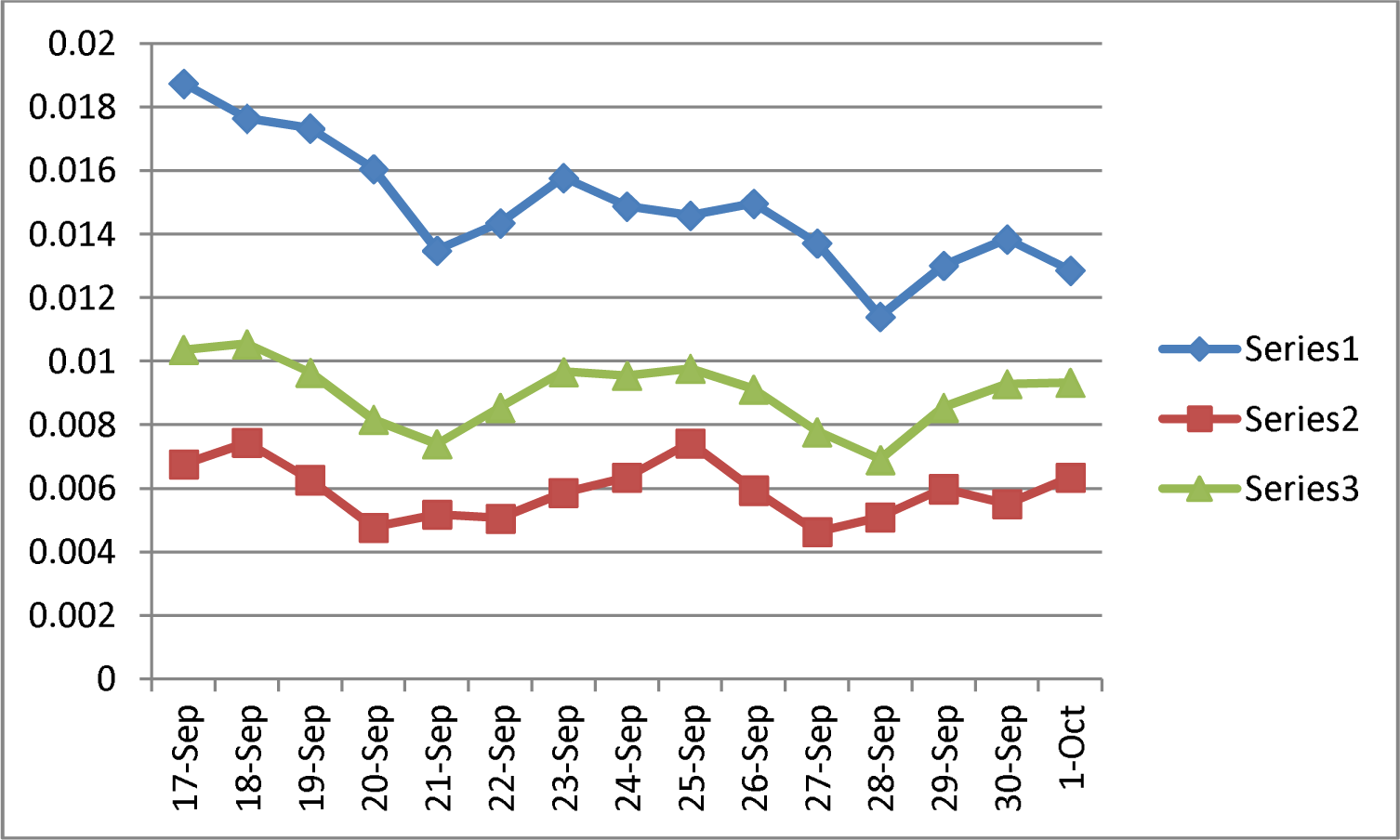
Values of Δ*z*(*t*): Series 1- India, Series 2- USA, Series 3- World.

From Fig, 1, we can clearly see the following things. In India, as the values of Δ*z*(*t*) are small enough and are still in a decreasing trend, the spread pattern is approximately exponential and that very soon the first order differences of *z*(*t*) would start to show approximate constancy, after which the situation in India would start improving. We can see further that the values of Δ*z*(*t*) in the case of the United States are very small and are very nearly constant, and the current exponential pattern would soon change to a nearly logarithmic pattern. In the case of the World as a whole, the smallness and the nearly constant values of Δ*z*(*t*) assert that the current exponential pattern of growth of the pandemic would soon change towards a nearly logarithmic pattern soon. We can say further that the growth would retard in the United States first to be followed by the World as a whole. Finally, when Δ*z*(*t*) would become very nearly constant in India which is currently the largest contributor towards the growth of the cumulative total number of cases, the situation in the World would automatically change towards a logarithmic increasing pattern which is the last phase of growth of a pandemic. A nearly logarithmic pattern of growth in the retarding phase of a pandemic we have observed in the case of Italy [10].

We now proceed to find the forecasts from October 2 to October 16 using the following method. The averages of Δ*z*(*t*) from September 17 to October 1 were found to be 0.008978 for the World, 0.005911 for the USA and 0.014846 for India.

We shall apply the following equation for forecasts of the cumulative total in the World

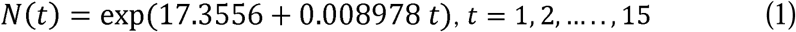

where exp(17.3556) = 34469967, the cumulative total number of cases in the World on October 1. This will give us the forecasts from October 2 onwards for the next 15 days.

For the United States, the equation we shall apply is

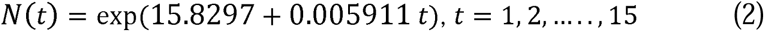

where exp(15.8297) = 7494671, the cumulative total number of cases in the United States on October 1. This will give us the forecasts from October 2 onwards for the next 15 days.

For India, the equation we shall apply is

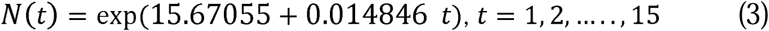

where exp(15.67055) = 6391960, the cumulative total number of cases in India on October 1. This will give us the forecasts from October 2 onwards for the next 15 days.

In Table-4, we have shown the forecasts for the World, the United States and for India. We would like to mention at this point that the forecasts may be overestimates for India in particular because the values of Δ*z*(*t*) were decreasing in the period from September 17 to October 1 and we have taken help of the average of Δ*z*(*t*) for the forecasts.

**Table-4:** Forecasts of the Cumulative Total Number of Cases.

| **Dates** | **World** | **USA** | **India** |
| --- | --- | --- | --- |
| 2-Oct | 34780848 | 7539103 | 6487563 |
| 3-Oct | 35094533 | 7583798 | 6584595 |
| 4-Oct | 35411047 | 7628759 | 6683079 |
| 5-Oct | 35730415 | 7673986 | 6783035 |
| 6-Oct | 36052664 | 7719481 | 6884487 |
| 7-Oct | 36377820 | 7765245 | 6987456 |
| 8-Oct | 36705907 | 7811281 | 7091965 |
| 9-Oct | 37036954 | 7857590 | 7198037 |
| 10-Oct | 37370987 | 7904174 | 7305696 |
| 11-Oct | 37708032 | 7951034 | 7414965 |
| 12-Oct | 38048116 | 7998171 | 7525868 |
| 13-Oct | 38391268 | 8045588 | 7638430 |
| 14-Oct | 38737515 | 8093286 | 7752676 |
| 15-Oct | 39086885 | 8141267 | 7868630 |
| 16-Oct | 39439406 | 8189532 | 7986319 |

## Conclusions

We conclude that the growth of the pandemic in the United States is in the threshold already to take a change from exponential to nearly logarithmic. As far as the situation in India is concerned, we can conclude that the pattern of growth is still approximately exponential and that very soon the situation in India would enter into the retarding phase of the pandemic. For the situation in the World as a whole too we can conclude that the growth of the pandemic is already in the threshold to take a change from exponential to logarithmic. However till India continues to be in the nearly exponential phase, the retardation in the World as a whole would have to wait.

## Data Availability

Worldometers.info

## REFERENCES

1. W. Kermak, A. McKendrick. (1927). A Contribution to the Mathematical Theory of Epidemics, Proceedings of the Royal Society A. Vol. 115 (1927): 700–72. (Republished as Contributions to the Mathematical Theory of Epidemics – I, Bulletin of the Mathematical Biology, (1991) 53 (1-2): 33–55.

2. W. Kermak, A. McKendrick. (1932) Contributions to the Mathematical Theory of Epidemics II. The Problem of Endemicity, Proceedings of the Royal Society A. Vol. 138: 55–83. (Republished in Bulletin of the Mathematical Biology, (1991) 53 (1-2): 57-87.

3. W. Kermak, A. McKendrick. Contributions to the Mathematical Theory of Epidemics III. Further Studies of the Problem of Endemicity, Proceedings of the Royal Society A. Vol. 141 (1933): 94–122. Bulletin of the Mathematical Biology, (1991) 53 (1-2): 89-118.

4. Worldometers.info. Total corona virus cases in India, Publishing Date: October 2 2020. Place of Publication: Dover, Delaware, U. S. A.

5. H. K. Baruah, A Simple Method of Finding an Approximate Pattern of the COVID-19 Spread, medRxiv preprint doi: https://doi.org/10.1101/2020.05.24.20112292. Posted on May 30, 2020.

6. H. K. Baruah, On Reliability of the COVID-19 Forecasts, medRxiv preprint doi: https://doi.org/10.1101/2020.06.01.20118844. Posted on June 1, 2020.

7. H. K. Baruah, The current COVID-19 spread pattern in India, medRxiv preprint doi: https://doi.org/10.1101/2020.06.03.20121210. Posted on June 8, 2020.

8. H. K. Baruah, Nearly Perfect Forecasting of the Total COVID-19 Cases in India: A Numerical Approach, medRxiv preprint doi: https://doi.org/10.1101/2020.06.13.20130096. Posted on June 13, 2020.

9. H. K. Baruah, The Uncertain COVID-19 Spread Pattern in India: A Statistical Analysis of the Current Situation, medRxiv preprint doi: https://doi.org/10.1101/2020.08.30.20184598. Posted on September 2, 2020.

10. H. K. Baruah, The Covid-19 Spread Patterns in Italy and India: A Comparison of the Current Situations, medRxiv preprint doi: https://doi.org/10.1101/2020.06.21.20136630. Posted on June 23, 2020.

